# Leveraging questionnaire-based physical activity levels (PAL) to identify energy intake misreporting using the Goldberg method: A doubly labeled water validation study

**DOI:** 10.1101/2025.04.02.25325112

**Authors:** Heather K Neilson, Shervin Asgari, Janet A. Tooze, Farah Khandwala, Anita Koushik, Rémi Rabasa-Lhoret, Karen A. Kopciuk, Ilona Csizmadi

**Author notes:** Corresponding Author: Dr Ilona Csizmadi, Departments of Community Health Sciences, Cumming School of Medicine, University of Calgary, Calgary, Alberta, Canada. Co-senior authors (Karen A. Kopciuk, Ilona Csizmadi). **Sources of Support:** Funding for this study was provided to institutions by the Canadian Institutes of Health Research (Grant MOP-86632) and the Alberta Cancer Research Institute (Grant 24265). **Role of funding agencies:** The funders had no role in the design of this study, the analysis, interpretation of results or drafting of the manuscript.

## Abstract

The Goldberg method has been suggested for identifying energy intake (EI) under-reporting in nutritional epidemiology. Its implementation, however, is limited by challenges associated with estimating physical activity levels (PAL). We quantified the accuracy of the Sedentary Time and Activity Reporting Questionnaire (STAR-Q) derived PAL (PAL_STAR-Q_) combined with the Goldberg method (Goldberg-PAL_STARQ_) to identify EI misreporting as compared with doubly labeled water (DLW) derived total energy expenditure (TEE_DLW_). Between 2009 and 2011, 99 men and women completed a two-week DLW protocol, a food frequency questionnaire, and the STAR-Q. The sensitivity, specificity, positive predictive value (PPV), negative predictive value (NPV) and accuracy of the Goldberg-PAL_STAR-Q_ were determined. Fifty-eight percent of men and women were classified as under-reporters by Goldberg-PAL_STAR-Q_ compared with 60% of men and 56% of women by TEE_DLW_. Among men, sensitivity, specificity, PPV, NPV and accuracy and 95% confidence intervals were 88% (68%-97%), 87% (61%-98%), 91% (72%-99%), 81% (56%-94%), and 87% (72%-95%), respectively; and among women 79% (62%-90%), 69% (50%-84%), 77% (60%-88%), 72% (52%-86%), and 75% (62%-84%), respectively. Validated individual level PALs used with the Goldberg method can be informative in sensitivity analyses to gain insight into EI misreporting in nutritional epidemiology studies lacking in objective EI measures.

## INTRODUCTION

Under-reporting of energy intake (EI) on dietary questonnaires is a widely recognized problem that distorts estimates of dietary intake in nutrition surveys and etiologic studies – thus impeding progress in diet-related research (1). Various statistical approaches have been suggested to identify dietary intake misreporting, but none have satisfactorily addressed the problem (2). Regression calibration of dietary intake is a promising approach for nutrients and dietary patterns (3–5); however, the method relies on the knowledge of *true* dietary intake and an understanding of the underlying systematic errors associated with self-report which are generally unknown (6). Ideally, misreporting could be detected using ‘gold standard’ objective biomarkers of dietary intake, but, these methods are limited in number and usually too labor-intensive and costly to feasibly implement in large-scale epidemiologic studies (7). A practical alternative, specifically for EI is the classification of individuals according to the plausibility of their reported EI (rEI) compared with their energy requirements (8).

The Goldberg method (9), originally proposed for identifying EI misreporting in community-dwelling populations, is based on the principle that under long-term, weight-stable conditions, EI is approximately equal to total energy expenditure (TEE). In addition, the method leverages the well-established relations between EI, TEE, basal metabolic rate (BMR) and physical activity level (PAL), where EI/BMR approximates TEE/BMR which in turn represents PAL (10, 11). Further, the method proposes the estimation of Goldberg cut-point boundaries, or 95% confidence intervals (95% CI), against which ratios of rEI:BMR are compared to identify under and over-reporting.

While conceptually tenable, the method is logistically challenged by difficulties associated with estimating PAL in free-living populations (11, 12). In the absence of population-specific objective measures, a low-activity PAL of 1.55 based on World Health Organization energy requirement prediction equations (13) was applied in the original Goldberg paper (14). The Goldberg method combined with a globally applied PAL of 1.55 was subsequently compared with doubly labelled water (DLW), the ‘gold standard’ for estimating TEE (15, 16), and reported to have acceptable accuracy in a study of middle-aged men and women; however, alternate PALs were not tested (17). More recently, contemporary DLW studies included in the report from the Dietary Reference Intake (DRI) for Energy (10), point to a median adult PAL of 1.68, suggesting that a global PAL of 1.55 may be unrealistically low for substantial segments of the population potentially resulting in very active under-reporters misclassified as acceptable.

To our knowledge, the Goldberg method has not been evaluated for accuracy using PALs derived from comprehensive individual self-reports of activity despite their value in capturing habitual behaviors of relevance in nutritional epidemiology. With that in mind, we appraised the accuracy of the Goldberg method for classifying EI reporting status using individual-level PALs derived from the Sedentary Time and Activity Reporting Questionnaire (STAR-Q), a comprehensive all-inclusive activity questionnnaire previously validated against DLW (18). In our primary analysis, we determined the accuracy of the Goldberg method using STAR-Q-derived PALs, hereafter referred to as Goldberg-PAL_STAR-Q_, in comparison with DLW-derived TEE_DLW_ for identifying EI misreporting on a food frequency questionnaire (FFQ). In secondary analyses we similarly tested the commonly used PAL of 1.55 and in sensitivity analyses we modified the assumption for FFQ variability in estimating of 95% CIs (17).

## METHODS

Participants in this analysis were those enrolled in the original STAR-Q DLW validation study (18). The STAR-Q was conceptualized to capture 24-hour movement behaviours (19). It was therefore uniquely designed to ascertain self-reported time spent in sleep, sedentary behavior, and all types and intensities (e.g., light, moderate and vigorous) and domains of activity (e.g., occupation, leisure, household, and transportation) and took a median time of 34 minutes to complete (18, 20).

### Subject Eligibility and Ethics

Participant recruitment has previously been described (18). Briefly, urban residents living in Calgary, Alberta, Canada were recruited from the community or from the Alberta Tomorrow Project (ATP). The ATP is a large geographically dispersed cohort established in 2001 in Alberta, to study links between lifestyle behaviors and chronic disease risk (21, 22). ATP participants who met eligibility criteria for the DLW validation study – body mass index (BMI) ≤ 35, weight stable (± 2.5 kg for at least 3 months), not planning to gain or lose weight, not pregnant or breastfeeding, not diagnosed with metabolic disorders, and not taking medications that modify water balance – were actively recruited by mail. Non-ATP participants were recruited by community-based advertisements. The upper age limit for eligibility was 60 years, and the lower age limit was 30 and 35 years for urban Calgary and ATP participants, respectively.

All participants provided written informed consent. Ethical approval was obtained from the Alberta Cancer Research Ethics Committee of Alberta Health Services and the Conjoint Health Research Ethics Board of the University of Calgary. All study procedures were in accordance with ethical standards outlined by the two governing bodies.

### Doubly labeled water study

Details of the 14-day DLW protocol have been published (18). Briefly, at baseline (Day 0), following an overnight fast, participants provided urine and saliva samples to determine background isotope levels. Each subject was then orally dosed with 2.5g 10 atom % of_18_ O per kg total body water (TBW) and 0.18 g 99 atom % deuterium (^2^H) per kg estimated TBW (Rotem Inc., Topsfield, Massachusetts). Post-dose saliva samples were collected at 3 and 4 hours to estimate TBW from deuterium-isotope dilution, and second-void urine samples on Days 1, 8, and 14. On Day 14 after an overnight fast, participants received 0.18 g 99 atom % ^2^H per kg TBW. Post-dose saliva sample collection was as described for Day 0. To estimate the decline in isotope enrichment, samples were batch-analyzed in duplicate using the Isoprime Stable Isotope Ratio Mass Spectrometer and the Multiflow-Bio module for Isoprime equipped with a Gilson 222XL Autosampler (GV Instruments, Manchester, United Kingdom). Data processing was performed using IonVantage (2012) software for Isoprime (Cheadle, United Kingdom).

We calculated TEE_DLW_ in kilocalories per day using a modified Weir equation (23) and an assumed respiratory quotient of 0.85. In the absence of objective measures, BMR was estimated using the Schofield Equation (24) to derive individual level PALs from DLW as:

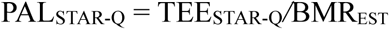

### Health and lifestyle measurements

Height and weight were measured (to the nearest 0.1 cm and 0.1 kg, respectively) on Days 0 and 14 of the DLW protocol using standard procedures (25). The TBF-310 Tanita Body Composition Analyzer and Scale (TheCompetitiveEdge.com, Preston, Washington) was used to measure body weight. BMI was determined as body weight divided by height-squared (kg/m^2^).

The Canadian Diet History Questionnaire I (C-DHQ I) was administered once on Day 0 of the DLW protocol to estimate rEI. The C-DHQ I is a comprehensive FFQ that assesses usual dietary intake over the past-year, modified from the U.S. National Cancer Institute’s DHQ for Canadian populations (26, 27) and used in large epidemiologic studies including the ATP (21, 22, 28, 29).

The STAR-Q was completed on Day 14 of the DLW study to estimate average daily energy expenditure from all activities including sleep during the preceding month. STAR-Q-derived TEE and activity energy expenditure demonstrated high reproducibility over a three-month period, with intraclass correlation coefficients of 0.84 and 0.73, respectively (18). Compared with DLW, the STAR-Q demonstrated moderate validity for ranking individuals by TEE and activity energy expenditure (Spearman’s rho=0.53 and 0.40, respectively) (18).

To derive TEE from the STAR-Q, activity code and metabolic equivalents of task (MET) (30) were first assigned to each reported activity. If the total time reported for physical activity, sedentary behavior, and sleep was less than 24 hours/day, a MET value of 1.2 was assigned for unaccounted time based on evidence that sedentary activity is more likely to be under-reported than other intensity levels of activity (18, 31). Reported frequency, duration and assigned MET values were used to estimate energy expenditure for each activity in units of MET-hours/day, which were then summed to derive the total average MET-hours/day. The STAR-Q-derived TEE (TEE_STAR-Q_) was then estimated as:

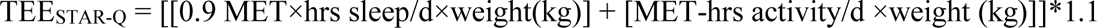

where STAR-Q reported hours of sleep per day was multiplied by 0.9 METs (30, 32) and then added to the total average MET-hours/day in awake time multiplied by baseline body weight. The equivalence of 1 MET = 1 kcal/kg/hr was applied to estimate the energy cost of activities (33) and 10% was added for the thermic effect of food (10).

Individual PALs were derived from self-report (TEE_STAR-Q_) as:

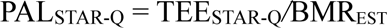

where BMR_EST_ was estimated using the Schofield sex-stratified equations based on age, weight and height (24).

### Classification of misreporting

EI reporting status was determined for each of the ratios rEI:MBR_EST_, rEI:TEE_DLW_, and rEI:TEE_STARQ_ compared with respective estimated 95% CIs based on PAL_STAR-Q_, TEE_DLW_, and TEE_STAR-Q_ (calculations below). Each participant was classified as an under-reporter (UR), acceptable reporter (AR), or over-reporter (OR) according to comparisons of individual rEI ratios with 95% CIs. Participants with ratios falling below their respective 95% CIs were deemed URs, those with ratios above the upper boundary were deemed ORs and those with ratios between the boundaries were deemed ARs.

In the primary analysis, to evaluate the Goldberg-PAL_STAR-Q_ for classifying individual misreporting, 95% CIs for comparison with rEI:BMR_EST_ were calculated as:

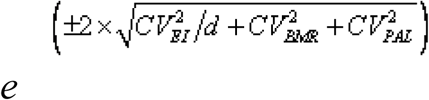

where CV_EI_ was the within-subject coefficient of variation (CV) for rEI, assigned as 18.6% for men and 19.8% for women based on repeatability of the US DHQ (17), *d* =1 since the C-DHQ I was administered once, CV_BMR_ was the CV for estimated BMR assumed to be 8.5% for men and women as previously suggested (11) and CV_PAL_ was the CV for PAL_STAR-Q_, calculated to be 17.5% for men and 15.1% for women from our three-month STAR-Q repeatability study (18).

For the gold standard rEI:TEE_DLW_ the 95% CIs were calculated as:

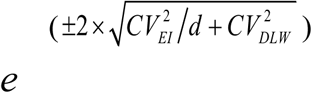

where CV_EI_ (*d* =1) was assumed to be the same as above and CV_DLW_ was assigned 5.1% for both men and women as previously reported (17).

To examine the magnitude of discrepancies relative to the gold standard ratio rEI:TEE_DLW_, graphical illustrations described by Tooze *et al*. (17) were used comparing individual-level rEI:TEE_STAR-Q_ and rEI:TEE_DLW_ ratios to their respective 95% CIs.

The 95% CI for with which rEI/TEE_STAR-Q_ was compared was calculated as:

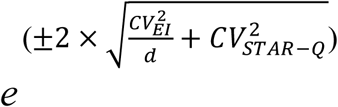

where CV_EI_ and *d* were as described above. The CV_STAR-Q_ was the within-subject CV for TEE_STAR-Q_ calculated to be 17.7% for men and 15.2% for women based on the first and second administrations of the STAR-Q in the repeatability study (18). If an individual rEI:TEE_STAR-Q_ ratio fell between the lower (or upper) 95% CI boundaries for rEI:TEE_STAR-Q_ and rEI:TEE_DLW_ ratios, the source of discrepancy was attributed to differences in the 95% CI calculations. Otherwise, it was attributed to differences in TEE estimation (TEE_STAR-Q_ vs TEE_DLW_). Furthermore, the contribution of these discrepancies to sensitivity and specificity of Goldberg-PAL_STAR-Q_ was determined.

In secondary analyses, a constant PAL 1.55 was assumed for participants (PAL_1.55_) and 95% CIs calculated as described above for PAL_STAR-Q_, except CV_PAL_ was assumed to be 15% for both genders as previously reported (11).

Finally in sensitivity analyses, for comparison with a previous study (17), *d*=∞ was substituted for *d*=1, in each of the cut-point equations (above) eliminating the CV^2^_EI_ term. This alternate estimate of ‘*d*’ assumes that the C-DHQ I ascertains habitual diet over an infinite time period.

### Statistics

All analyses were stratified by gender. Means (SD) were determined for continuous sociodemographic characteristics, frequencies and percentages for categorical characteristics, and medians (Q1, Q3) for rEI and energy expenditure. To appraise the validity of Goldberg-PAL_STAR-Q_ and PAL_1.55_ classifications, the estimated proportions of participants correctly classified as URs, ARs and ORs were compared with DLW classifications assumed to be *true* classifications. Sensitivity, specificity, positive predictive value (PPV), negative predictive value (NPV), and the accuracy of UR and AR classifications were estimated (34), excluding ORs identified by TEE_DLW,d=1_ (n=1) and TEE_DLW,d=∞_ (n=7). The sensitivity was the proportion of true URs correctly classified by the Goldberg method (PAL_STAR-Q_ or PAL_1.55_) and specificity was the proportion of true ARs correctly classified. PPV was the proportion of Goldberg method URs that were true URs, interpreted as the probability of being a true UR if classified as UR by Goldberg. NPV was the proportion of Goldberg ARs that were true ARs, interpreted as the probability of being a true AR if classified as AR by the Goldberg method. Accuracy was determined as the proportion of true URs plus true ARs classified correctly with the Goldberg method.

To appraise the validity of TEE_STAR-Q_ for estimating the magnitude of misreporting (UR and OR) in kilocalories per day, the within-person differences of TEE_STAR-Q_ - rEI were compared with TEE_DLW_ - rEI using Pearson correlation coefficients and Wilcoxon signed-rank tests.

Agresti Coull 95% CIs were calculated for diagnostic metrics using Python (Version 3.12.12; Statsmodels Version 0.14.5). All other analyses were performed using SAS Institute Software, Gary, NC (Version 9.4). A *P*-value < 0.05 was considered statistically significant.

## RESULTS

Participants aged 30-60 years (n=106) were recruited to the original DLW study between July 2009 and July 2010 (18). Seven participants were excluded from the present analysis for reasons outlined in **Figure 1**, leaving 99 participants of whom 60% were women. Compared with men, women were somewhat younger (mean: 46.2 vs 50.6 years of age), had a lower BMI (mean: 23.5 vs 25.9 kg/m^2^), and were less likely to have completed post-secondary education (45% vs 51%) (**Table 1)**. Most of the participants were Caucasian and had paid or volunteer work. Median EI, TEE_DLW_, TEE_STAR-Q_, and BMR_EST_ were lower for women (1676 kcal, 2604 kcal, 2889 kcal and 1362 kcal, respectively) compared with men (2036 kcal, 3306 kcal, 3620 kcal, and 1866 kcal, respectively). Median values for PAL_DLW_ and PAL_STAR-Q_, were 1.8 and 2.0 for men and, 1.9 and 2.1 for women **(Table 1)**, consistent with ‘active’ and ‘very active’ PALs (10).

**Figure 1.**
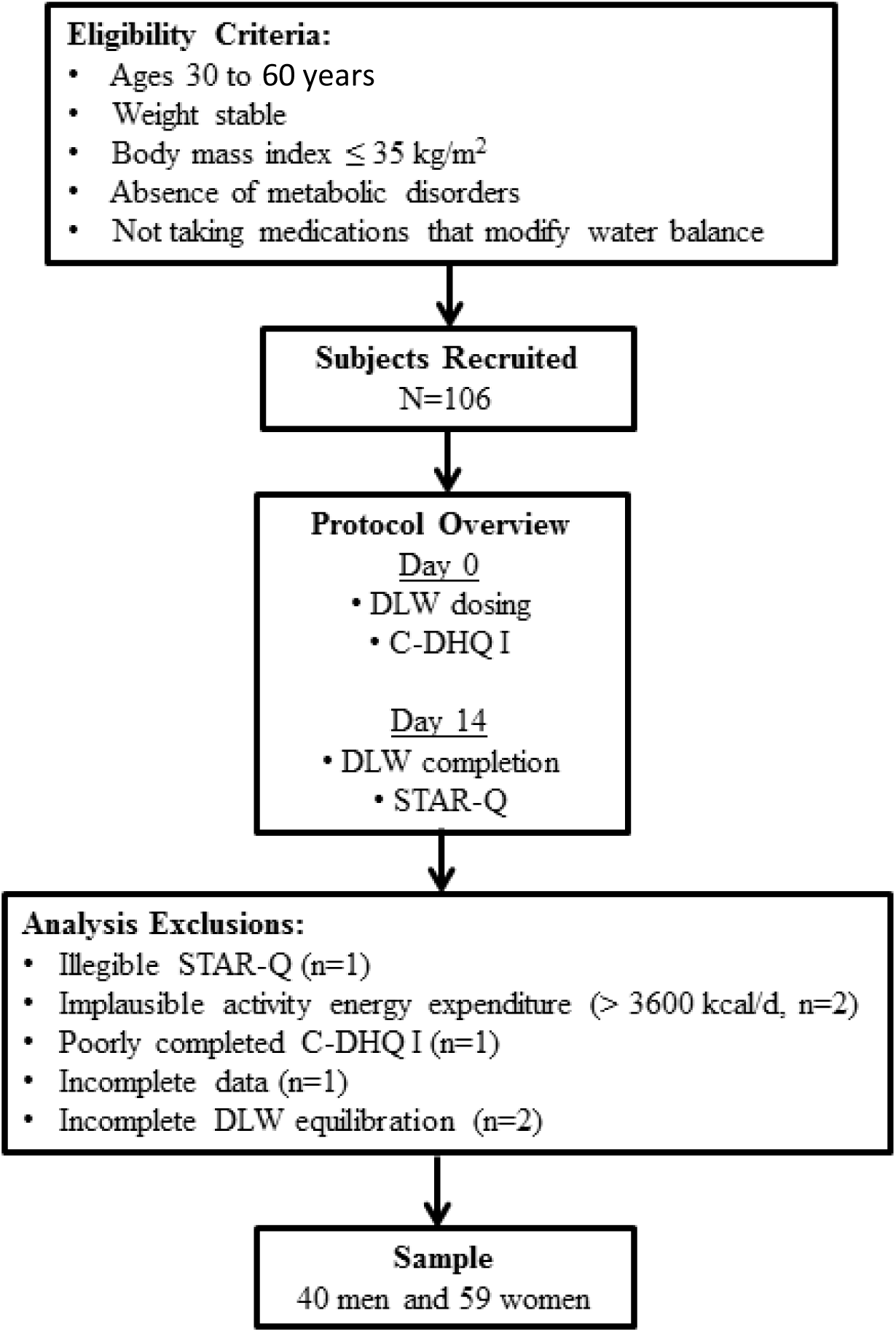
Participant recruitment in the STAR-Q validation study (2009-2011), Alberta, Canada.

**Table 1.** Sociodemographic characteristics, energy expenditure and energy intake for 99 participants in the STAR-Q validation study. (**2009-2011**)**, Alberta, Canada.**

| Sociodemographic characteristics | Men, *n* = 40 |  | Women, *n* = 59 |  |
| --- | --- | --- | --- | --- |
|  | Mean (SD) | % | Mean (SD) | % |
| Age, years | 50.6 (7.0) |  | 46.2 (8.5) |  |
| Body mass index, kg/m<sup>2</sup> | 25.9 (3.1) |  | 23.5 (2.9) |  |
| Education |  |  |  |  |
| Less than post-secondary education |  | 34 |  | 45 |
| At least post-secondary education |  | 51 |  | 45 |
| Missing |  | 15 |  | 10 |
| Ethnicity |  |  |  |  |
| Caucasian |  | 73 |  | 85 |
| Asian |  | 3 |  | 5 |
| Other |  | 3 |  | - |
| Missing |  | 20 |  | 10 |
| Paid or volunteer work |  | 93 |  | 93 |
| Energy intake and expenditure | Median [Q1, Q3] |  | Median [Q1, Q3] |  |
| Energy intake, kcal/d | 2036 [1627, 2634] |  | 1676 [1317, 2033] |  |
| Self-reported activity (STAR-Q), MET-h/d<sup>1</sup> | 28.8 [24.6, 32.6] |  | 30.6 [25.6, 39.5] |  |
| TEE<sub>DLW</sub>, kcal/d | 3306 [2843, 3727] |  | 2604 [2226, 2854] |  |
| TEE<sub>STAR-Q</sub>, kcal/d | 3620 [3178, 4019] |  | 2889 [2420, 3387] |  |
| BMR<sub>EST</sub>, kcal/d | 1866 [1744, 1924] |  | 1362 [1331, 1438] |  |
| PAL (TEE<sub>DLW</sub>:BMR<sub>EST</sub>) | 1.8 [1.6, 2.0] |  | 1.9 [1.7, 2.1] |  |
| PAL (TEE<sub>STAR-Q</sub>:BMR<sub>EST</sub>) | 2.0 [1.8, 2.1] |  | 2.1 [1.8, 2.4] |  |

<sup>1</sup> The sum of MET-h/d from all types of physical activity including light, moderate, and vigorous intensity and sedentary activities (sleep is excluded).
BMR<sub>EST</sub>, estimated basal metabolic rate; MET, metabolic equivalent of task; PAL, physical activity level; STAR-Q, Sedentary Time and Activity Reporting Questionnaire; TEE, total energy expenditure.

Results from the primary, secondary and sensitivity analyses are presented in **Table 2**. In the primary analysis, 60% of men and 56% of women were classified as URs by TEE_DLW_, and 58% of men and women by the Goldberg-PAL_STAR-Q_. The Goldberg-PAL_STAR-Q_ misclassified fewer men (15%) than women (25%), resulting in sensitivity, specificity, PPV, NPV and accuracy >80% among men, whereas for women these estimates were <80%.

**Table 2.**
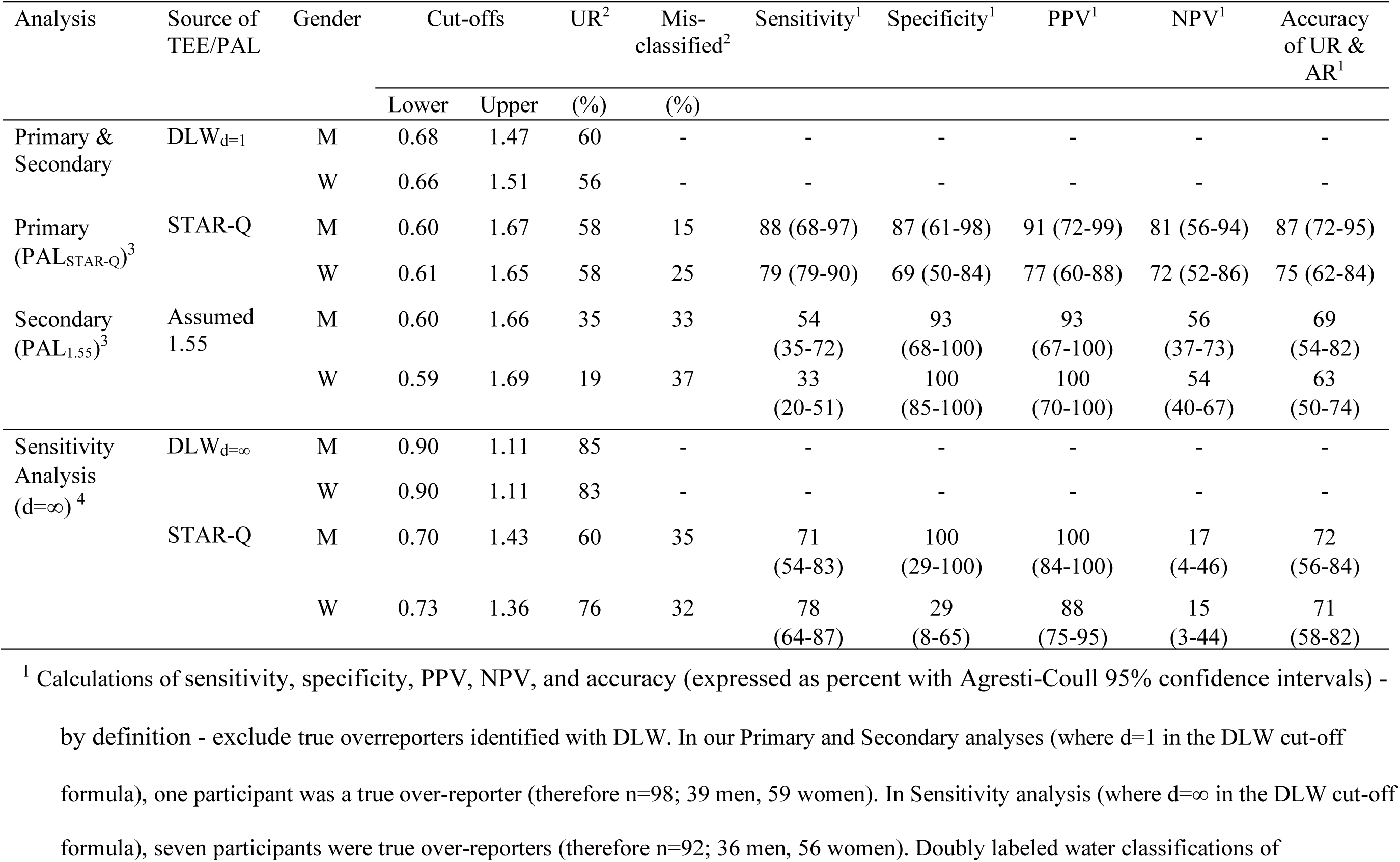
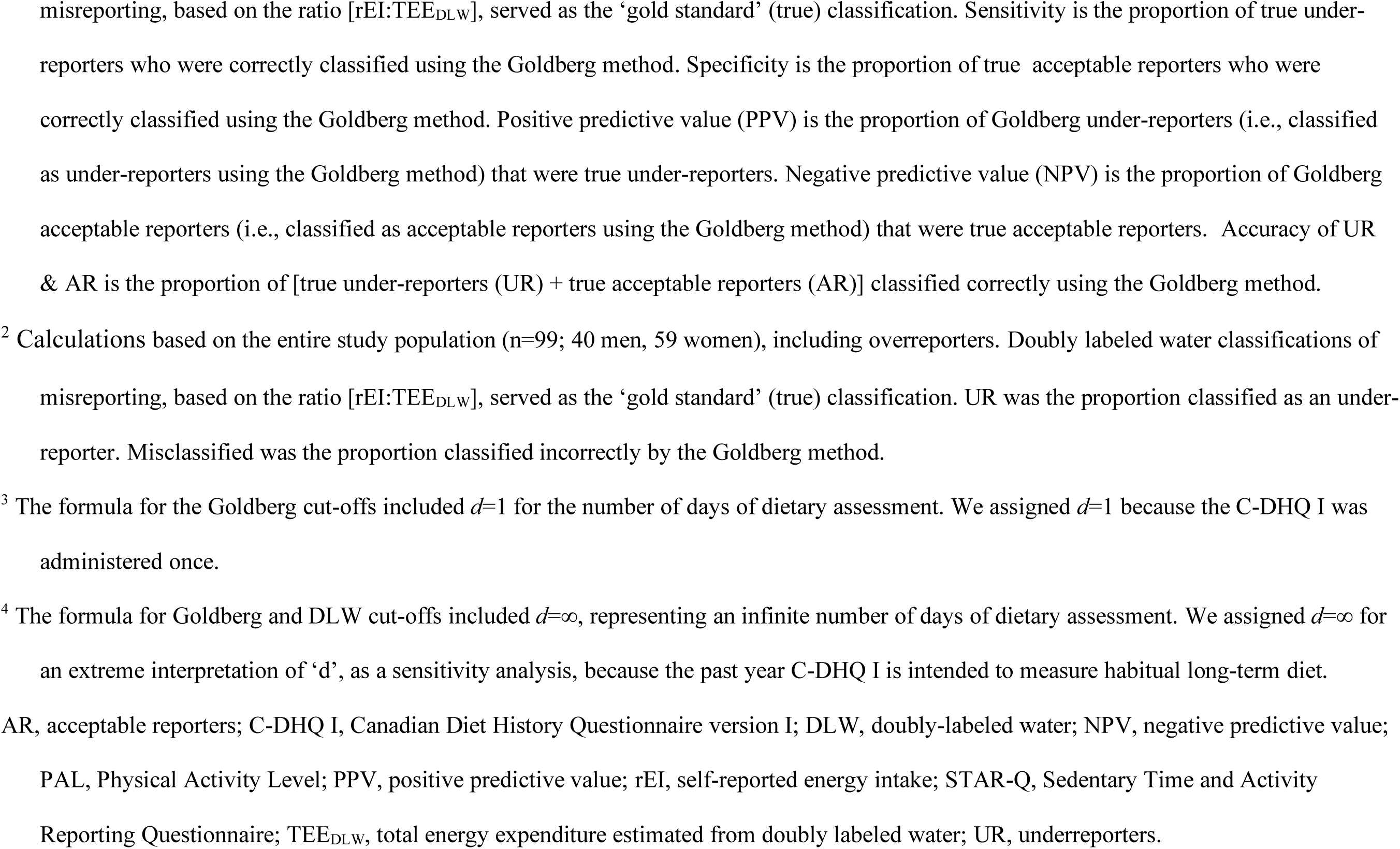
Validity of the Goldberg method compared with the ‘gold standard’ doubly labeled water method for classifying energy intake misreporting, for 40 men and 59 women^1^ in the STAR-Q validation study (2009-2011), Alberta, Canada.

| Analysis | Source of TEE/PAL | Gender | Cut-offs |  | UR<sup>2</sup> | Mis-classified<sup>2</sup> | Sensitivity<sup>1</sup> | Specificity<sup>1</sup> | PPV<sup>1</sup> | NPV<sup>1</sup> | Accuracy of UR & AR<sup>1</sup> |
| --- | --- | --- | --- | --- | --- | --- | --- | --- | --- | --- | --- |
|  |  |  | Lower | Upper | (%) | (%) |  |  |  |  |  |
| Primary & Secondary | DLW<sub>d=1</sub> | M | 0.68 | 1.47 | 60 | - | - | - | - | - | - |
|  |  | W | 0.66 | 1.51 | 56 | - | - | - | - | - | - |
| Primary (PAL<sub>STAR-Q</sub>)<sup>3</sup> | STAR-Q | M | 0.60 | 1.67 | 58 | 15 | 88 (68-97) | 87 (61-98) | 91 (72-99) | 81 (56-94) | 87 (72-95) |
|  |  | W | 0.61 | 1.65 | 58 | 25 | 79 (79-90) | 69 (50-84) | 77 (60-88) | 72 (52-86) | 75 (62-84) |
| Secondary (PAL<sub>1.55</sub>)<sup>3</sup> | Assumed 1.55 | M | 0.60 | 1.66 | 35 | 33 | 54 (35-72) | 93 (68-100) | 93 (67-100) | 56 (37-73) | 69 (54-82) |
|  |  | W | 0.59 | 1.69 | 19 | 37 | 33 (20-51) | 100 (85-100) | 100 (70-100) | 54 (40-67) | 63 (50-74) |
| Sensitivity Analysis (d=∞)<sup>4</sup> | DLW<sub>d=∞</sub> | M | 0.90 | 1.11 | 85 | - | - | - | - | - | - |
|  |  | W | 0.90 | 1.11 | 83 | - | - | - | - | - | - |
|  | STAR-Q | M | 0.70 | 1.43 | 60 | 35 | 71 (54-83) | 100 (29-100) | 100 (84-100) | 17 (4-46) | 72 (56-84) |
|  |  | W | 0.73 | 1.36 | 76 | 32 | 78 (64-87) | 29 (8-65) | 88 (75-95) | 15 (3-44) | 71 (58-82) |

<sup>1</sup> Calculations of sensitivity, specificity, PPV, NPV, and accuracy (expressed as percent with Agresti-Coull 95% confidence intervals) -
by definition - exclude true overreporters identified with DLW. In our Primary and Secondary analyses (where d=1 in the DLW cut-off formula), one participant was a true over-reporter (therefore n=98; 39 men, 59 women). In Sensitivity analysis (where d=∞ in the DLW cut-off formula), seven participants were true over-reporters (therefore n=92; 36 men, 56 women). Doubly labeled water classifications of
misreporting, based on the ratio $[rEI:TEE_{DLW}]$ , served as the ‘gold standard’ (true) classification. Sensitivity is the proportion of true under-reporters who were correctly classified using the Goldberg method. Specificity is the proportion of true acceptable reporters who were correctly classified using the Goldberg method. Positive predictive value (PPV) is the proportion of Goldberg under-reporters (i.e., classified as under-reporters using the Goldberg method) that were true under-reporters. Negative predictive value (NPV) is the proportion of Goldberg acceptable reporters (i.e., classified as acceptable reporters using the Goldberg method) that were true acceptable reporters. Accuracy of UR & AR is the proportion of [true under-reporters (UR) + true acceptable reporters (AR)] classified correctly using the Goldberg method.
<sup>2</sup> Calculations based on the entire study population (n=99; 40 men, 59 women), including overreporters. Doubly labeled water classifications of misreporting, based on the ratio $[rEI:TEE_{DLW}]$ , served as the ‘gold standard’ (true) classification. UR was the proportion classified as an under-reporter. Misclassified was the proportion classified incorrectly by the Goldberg method.
<sup>3</sup> The formula for the Goldberg cut-offs included $d=1$ for the number of days of dietary assessment. We assigned $d=1$ because the C-DHQ I was administered once.
<sup>4</sup> The formula for Goldberg and DLW cut-offs included $d=\infty$ , representing an infinite number of days of dietary assessment. We assigned $d=\infty$ for an extreme interpretation of ‘d’, as a sensitivity analysis, because the past year C-DHQ I is intended to measure habitual long-term diet.
AR, acceptable reporters; C-DHQ I, Canadian Diet History Questionnaire version I; DLW, doubly-labeled water; NPV, negative predictive value; PAL, Physical Activity Level; PPV, positive predictive value; rEI, self-reported energy intake; STAR-Q, Sedentary Time and Activity Reporting Questionnaire; $TEE_{DLW}$ , total energy expenditure estimated from doubly labeled water; UR, underreporters.

In secondary analyses, Goldberg-PAL_1.55_ resulted in fewer participants classified as URs (35% and 19% for men and women, respectively) than with TEE_DLW_ and Goldberg-PAL_STAR-Q_ which led to high specificity and PPVs (both 93% for men and 100% for women), but low sensitivity and NPVs (all <60% for men and women).

In sensitivity analyses, where *d*=∞ was assumed for the FFQ, a larger proportion of participants were classified as URs with both TEE_DLW_ and Goldberg-PAL_STAR-Q_ than with the assumption that *d*=1. Seven were deemed to be true ORs in contrast to one, when *d=*1, attributable to a narrower 95% CI with *d*=∞ following elimination of the variability term. As such, less favorable results were seen for the identification of URs with the Goldberg-PAL_STAR-Q_ except for specificity and PPV, which were 100% among men. However, the assumption that the FFQ captures habitual intake representing an infinite time period may be an overestimate of this attribute (35).

**Figures 2A and 2B** illustrate individual-level agreement and magnitude of misreporting in kilocalories per day (i.e., TEE_STAR-Q_ - rEI relative to TEE_DLW_ - rEI). Pearson correlation coefficients were higher for men (r=0.76, *P*<0.0001; classification agreement in 34 of 40 men) than for women (r=0.37, *P*=0.0042; classification agreement in 44 of 59 women). Nonetheless, the Wilcoxon signed-rank test indicated statistically significant within-person differences between the two methods (P = 0.0021 and P < 0.0001 for men and women, respectively). The magnitude of EI under-reporting tended to be larger for misclassified URs (e.g., URs classified as ARs, approximately 1,000 to 1,500 kcal/d; 3 men and 7 women) while the magnitude of EI under-reporting was typically smaller for misclassified ARs (e.g., ARs classified as URs, <1,000 kcal/d approximately; 2 men and 8 women).

**Figure 2.**
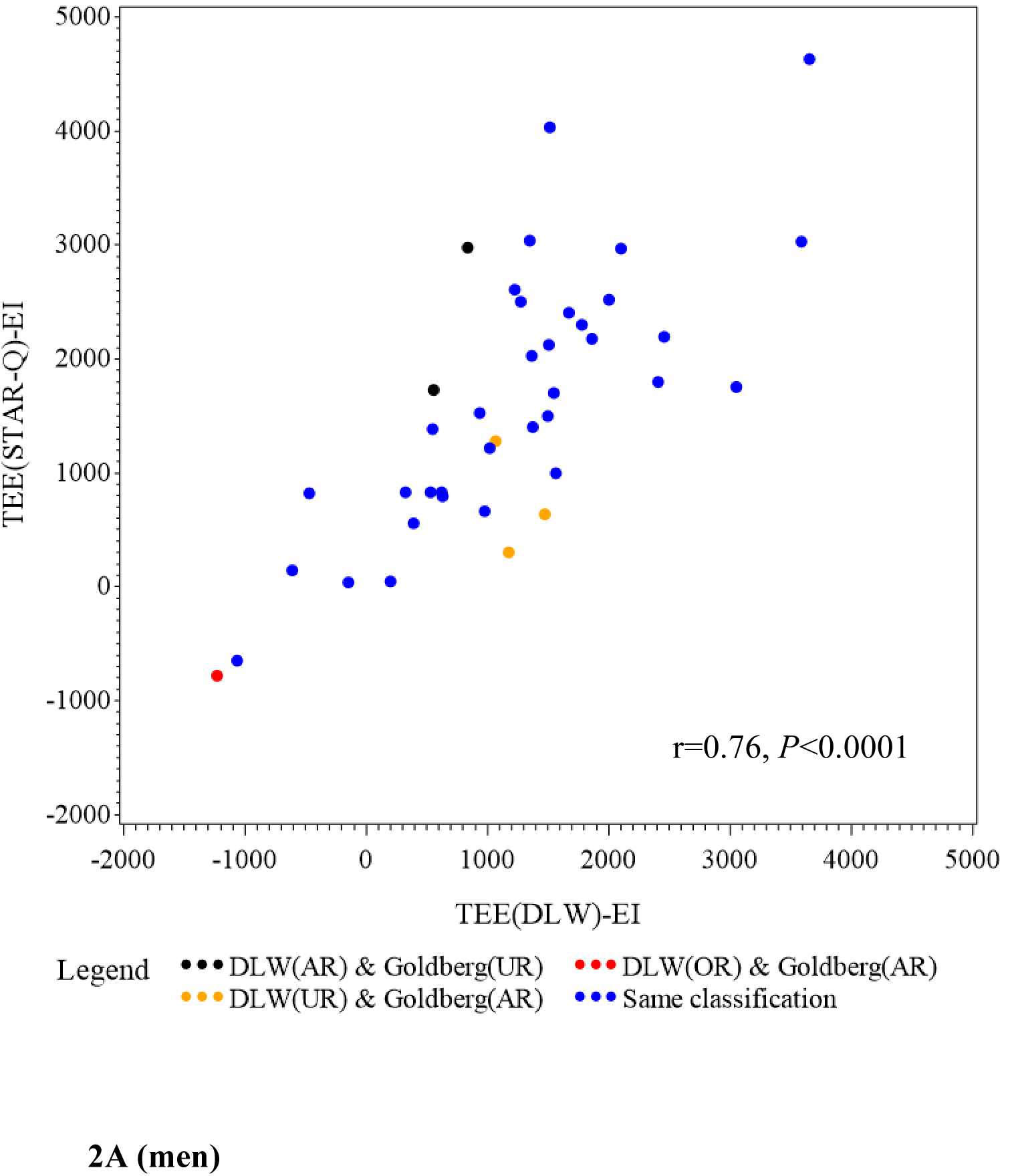
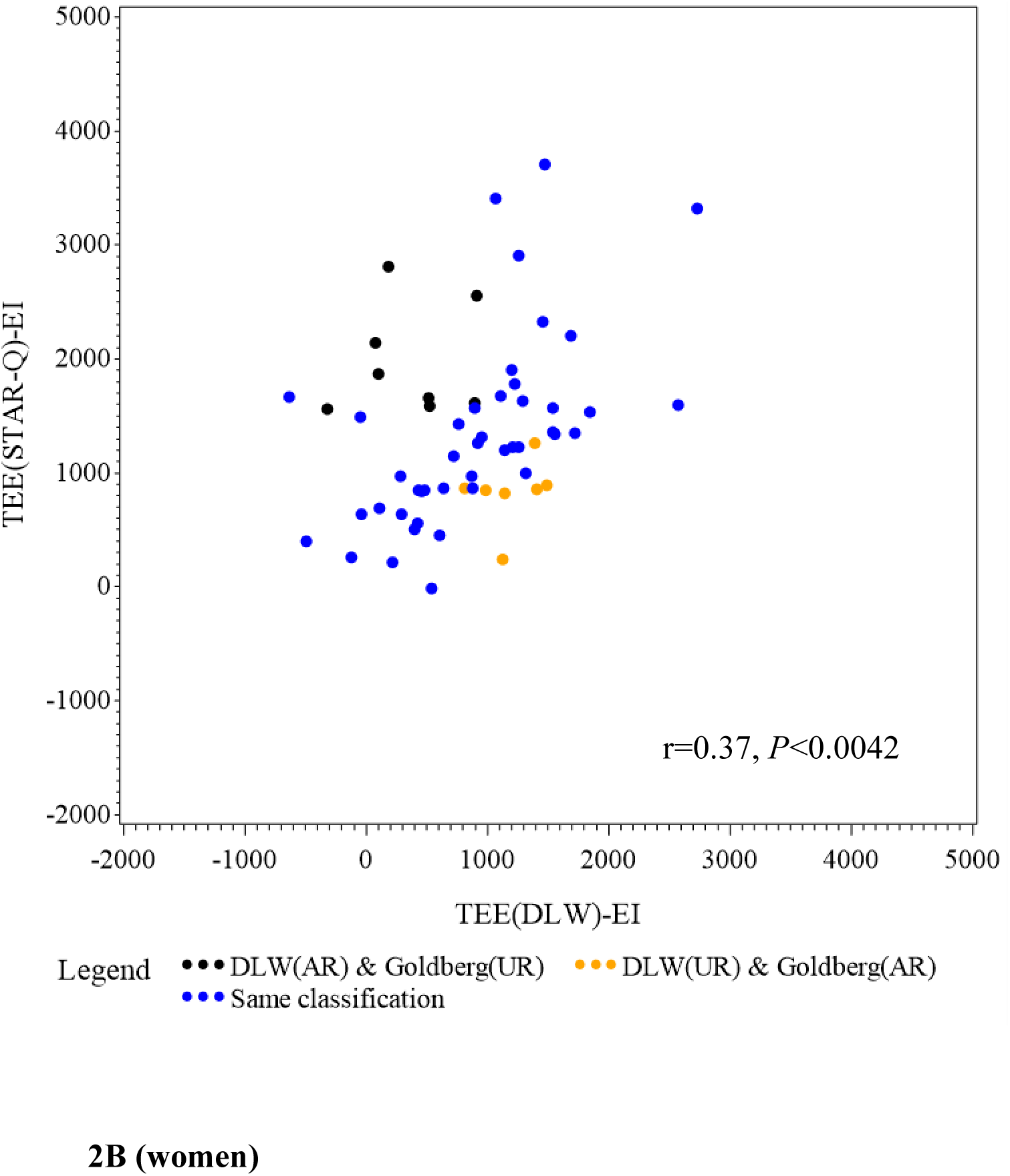
Correlation between the degree of energy intake misreporting based on Goldberg classifications (i.e., individual differences: TEE_STAR-Q_ – rEI) and the degree of energy intake misreporting based on doubly labeled water classifications (i.e., individual differences: TEE_DLW_-rEI) in kilocalories per day. **Figure 2A** shows the correlation among 40 men and **Figure 2B** shows the correlation among 59 women in the STAR-Q validation study (2009-2011), Alberta, Canada. AR, acceptable reporters; DLW, doubly labeled water; UR, under-reporters.

**Figures 3A and 3B**, show whether discrepant classifications arose from different 95% CI cut-points or from different TEE estimation. Among 40 men **(Figure 3A)**, five misclassifications were due to TEE estimation, and one was due to interval cut-point. Among 59 women (**Figure 3B)**, 11 misclassifications were due to differences in TEE estimation and 4 due to cut-point estimation. Among *true* URs misclassified as ARs using the PAL_STAR-Q_ (reflecting sensitivity), 2 of 3 men and 3 of 7 women were misclassified owing to TEE estimation. Among *true* ARs misclassified as URs using the PAL_STAR-Q_ method (reflecting specificity), all misclassifications (2 men and 8 women) were due to differences in TEE estimation.

**Figure 3.**
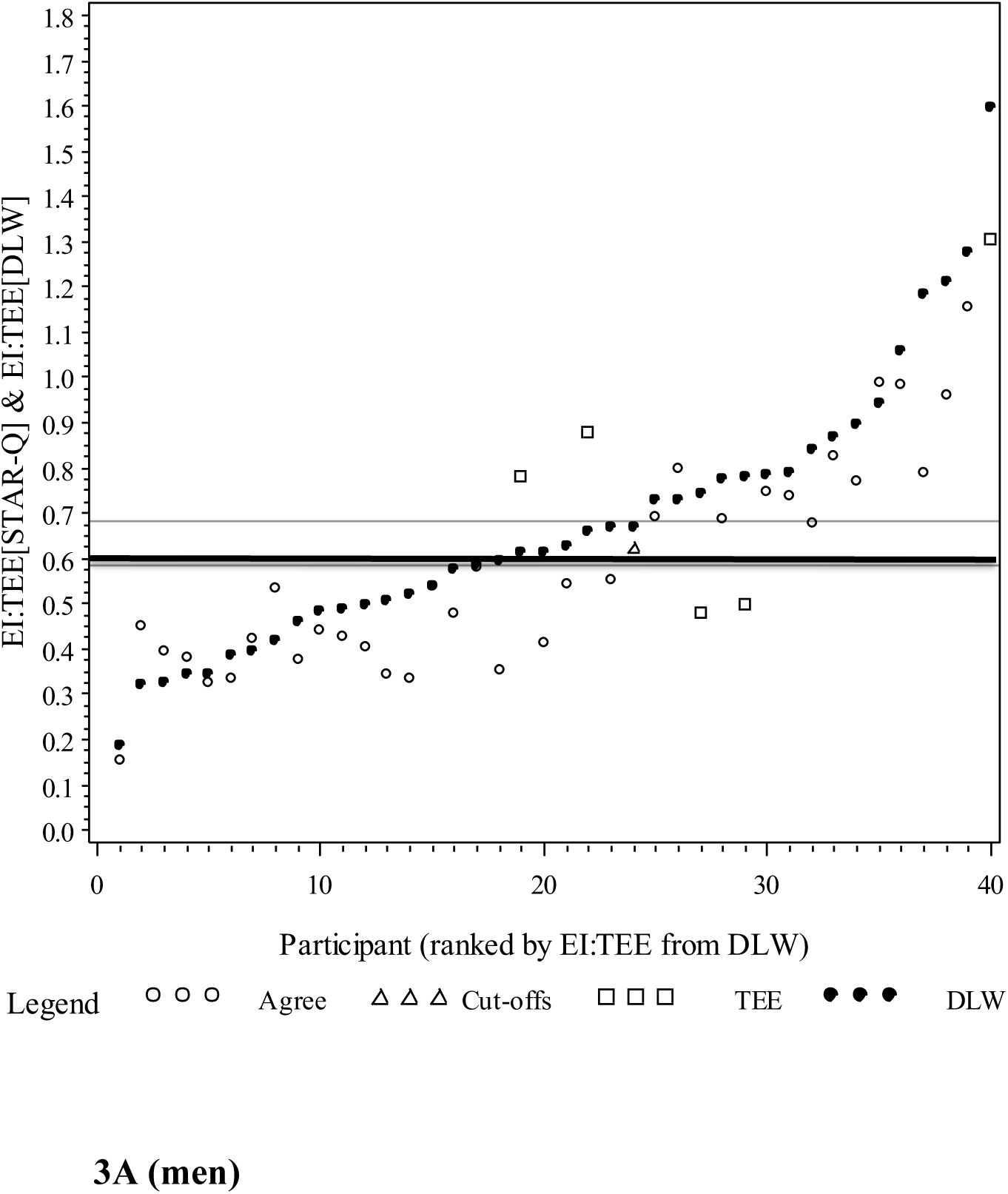
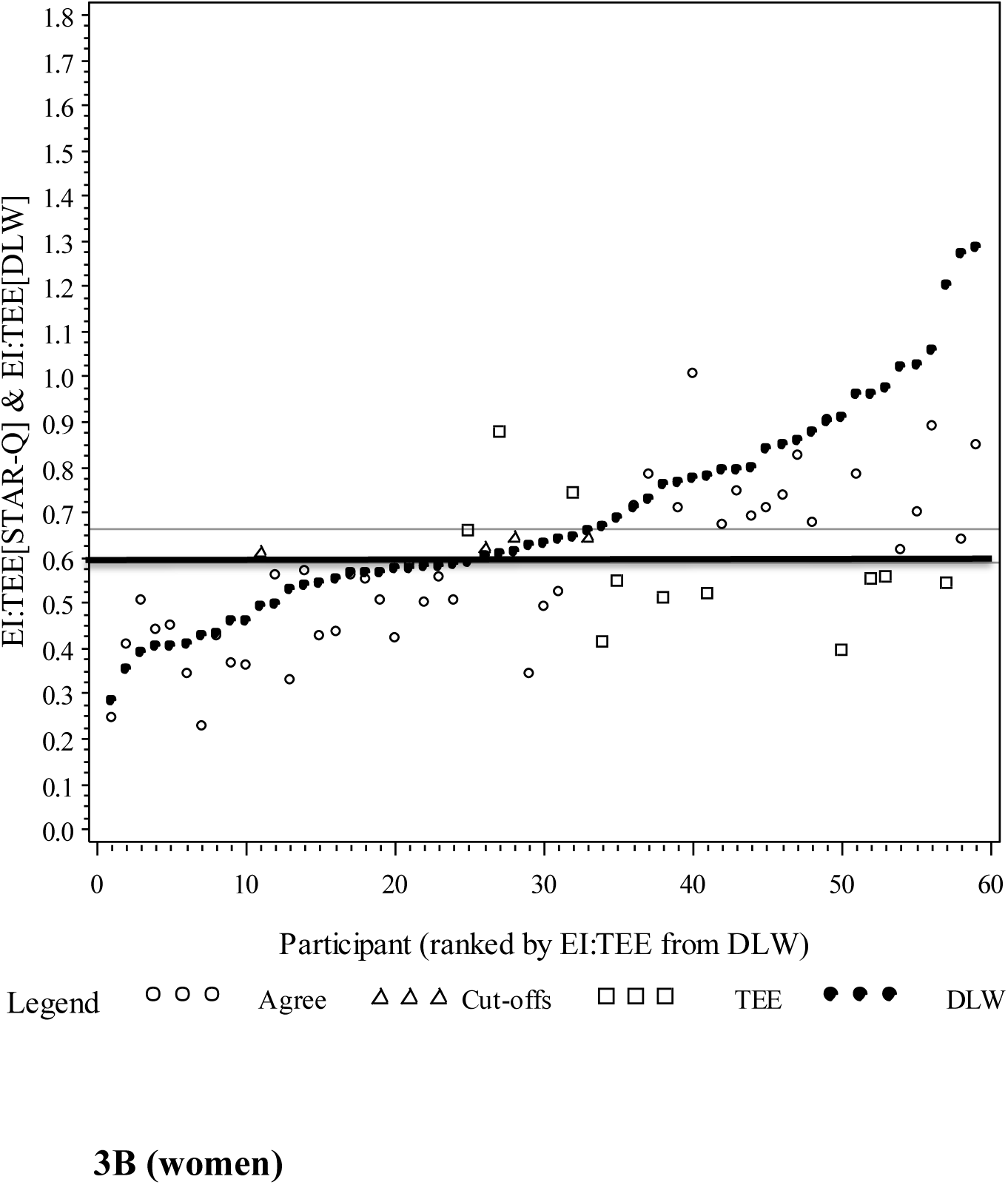
Sources of discrepancy between the Goldberg and doubly labeled water methods for classifying energy intake misreporting. **Figure 3A** illustrates sources of discrepancy in men and **Figure 3B** illustrates sources of discrepancy in women. The gold standard doubly labeled water classifications of reporting are represented with shaded circles rEI:TEE_DLW_. The Goldberg classifications of misreporting, based on the ratio rEI:TEE_STAR-Q_, are represented with open circles (agreement between methods), squares (discrepant because of TEE estimation) or triangles (discrepant because of different cut-offs). The gray solid line represents the lower cut-off for gold standard doubly labeled water classifications (0.68 for men; 0.66 for women). The black solid line represents the lower cut-off for Goldberg classifications (0.58 for men; 0.59 for women).

## DISCUSSION

Using DLW, the ‘gold-standard’ biomarker for TEE and an unbiased proxy for habitual EI in weight-stable individuals (2), we determined the accuracy of the Goldberg method using STAR-Q-derived PALs to classify EI reporting status among community dwelling middle-aged men and women following completion of an FFQ. The PAL_STAR-Q_ generally improved the sensitivity and overall accuracy of the Goldberg method for discriminating between URs and ARs compared with a global PAL of 1.55. We also identified gender-based differences in the performance of the Goldberg-PAL_STAR-Q_. In men, the method discriminated between UR and AR with an overall high level of accuracy at 87%, while in women it exhibited fair performance, with an overall classification accuracy of 75%.

A key determinant of the accuracy of the Goldberg method is the degree to which PALs represent populations under study. Three previous DLW validation studies have explored approaches to identify the optimal PAL for estimating EI misreporting (17, 36, 37). Two studies examined EI misreporting on food records and/or diet histories and found under-reporting in 14% (37) and 34% (36) of adult participants. Consistent with our results, the studies reported high levels of specificity but relatively low sensitivity for identifying URs when a global PAL of 1.55 was assumed. Additionally, Black (36) leveraged the strength of a large database comprised of 21 DLW studies (n=429) to test the effect of various global mean activity PALs (from 1.55 to 1.95) and reported incremental increases in sensitivity, with increasing PALs; however, this improvement coincided with decreases in specificity. In a small sample of adults (n=14), Livingstone *et al* (37) tested the performance of gender-specific PALs to detect URs based on individual heart-rate monitors matched to low, medium and high activity categories of PAL (13). Compared with PAL_1.55_, sensitivity increased substantially from 50% to 100% with negligible decreases in specificity. Subsequently, Tooze *et al* (17) reported favorable results for the Goldberg method with a PAL_1.55_ to detect DLW-classified URs on 24-hour dietary recalls (∼20%) and an FFQ (∼50% UR) in the Observing Protein and Energy Nutrition (OPEN) Study (n=451). Specificity was 88% for the FFQ in both genders. The sensitivity to detect URs on the FFQ was high at 96% when usual intake was assumed for the FFQ (*d=∞*), but substantially lower when the OPEN study FFQ within-person variability was applied (72% sensitivity for men and 62% for women).

The results reported by Livingstone *et al* (37) and Black (36) are consistent with our results and support the premise that a global PAL_1.55_ may be underestimating the activity levels of some adult populations – and hence, more individualized PALs may improve the overall accuracy of the Goldberg method to identify EI under-reporting. While our findings contrast those of Tooze *et al* (17), we note that the prevalence of true UR (50%) was lower in the OPEN study than in our population which affects PPV and NPV values. In addition, participants were on average less active and slightly older than in the STAR-Q validation study. Median values for DLW-derived TEE and PAL were 2,813 kcal and 1.69 for men and 2,283 kcal and 1.75 for women in OPEN (38, 39) and 3,306 kcal and 1.8 for men and 2604 kcal and 1.9 for women in the STAR-Q study, with ages ranging from 40 to 69 years in OPEN (38, 39) versus 30 to 60 years in the STAR-Q study.

Statistical approaches, such as the exclusion of misreporters, and stratification and adjustment for reporting status in regression analyses (40–45), have been explored to handle identified URs. Consensus exists only with respect to an agreement that exclusion of misreporters is undesirable given loss of statistical power (41) and valuable information inherent in excluded data (40). While the optimal approach may be study-specific (e.g., health outcomes of interest, study design, method of data collection and population characteristics), corrective methods cannot be expected to fully eliminate bias associated with measurement error. Nonetheless, the identification of implausible EI reporting using the Goldberg method has been demonstrated to significantly reduce, albeit not completely eliminate, misreporting-related bias for some EI-outcome associations (44, 45). This suggests an important role for the method in sensitivity analyses aiming to better understand the potential impact of EI misreporting on study results. Of note, the aforementioned studies used group level PALs – potentially limiting the extent to which bias due to misreporting could be mitigated.

While the STAR-Q was designed to capture a comprehensive profile of daily activities over 24-hours, the misclassification of 50% of true URs as ARs and all true ARs as URs, was likely due to errors in the estimation of TEE_STAR-Q_ **(Figure 3a and 3b)**. Using updated MET values (33, 46) for energy expenditure, could improve future TEE and PAL estimations. In addition, the reporting of activities during the previous month are not without cognitive challenges, though less so than past-year reporting (20). The Activities Completed over Time in 24 hours (ACT24), an internet-based recall of previous day activities is even less cognitively challenging and has been shown to estimate mean TEE and PAL values within 5% of DLW-based estimates (47, 48). The accuracy of short-term recall combined with longer-term measures that reflect habitual lifestyle behaviors is an intriguing approach that has been suggested for improving dietary intake measures (49) and may also improve TEE and PAL estimations. Moreover, new wearable device-based technologies may in the foreseeable future accurately estimate PALs and TEE in large populations, to complement the contextual data collected by traditional questionnaires (50). Hence, it is anticipated that methodologies combining device-derived estimates of energy expenditure and body composition may be developed to validate self-reported EI (51–53); though these approaches are still viewed as ‘works in progress’ (54).

A noteworthy strength of this study is the validation of the Goldberg-PAL_STAR-Q_ method against DLW – and the unique use of individual-level PALs not previously studied. Furthermore, as noted in the recent DRIs for Energy report, of 128 DLW studies representing 25 countries in the International Atomic Energy Agency DLW Database, none were Canadian (10) highlighting a gap in this area of Canadian research that this study begins to address. Nonetheless, our study has limitations that merit consideration. First, we used prediction equations for BMR (24), which accounts for a substantial proportion of TEE (55) and could be a source of error in the estimation of PAL_STAR-Q_. Second, we cannot be certain that some of the under-reporting was not true undereating, though we aimed to minimize this occurrence by recruiting generally weight stable participants to the DLW study. Third, we caution that we cannot assume that EI misreporting is reflective of misreporting of other nutrients. Indeed, though still relatively under-studied, evidence is accumulating to suggest that foods and nutrients vary in the accuracy with which they are reported (6, 56–59). On a final note, our population was largely Caucasian middle-aged adults; hence, our results may not be generalizable to older (60, 61), racially diverse populations and those with other characteristics that may affect reporting and health status (62–65).

Overall, our DLW-validated results lend credibility to the thesis that detailed comprehensive questionnaire-based activities may be effectively employed with the Goldberg method to describe and explore EI misreporting in large-scale diet studies. Importantly, such strategies could create opportunities to carry out sensitivity analyses that provide critical insights into the impact of EI misreporting on diet-disease associations in nutritional epidemiology. Ongoing effort to improve upon this approach with cohort validation sub-studies and innovative study designs integrated with new technologies need to remain as research priorities in nutritional epidemiology.

## STATEMENTS AND DECLARATIONS

### Data availability

Data described in the manuscript, code book, and analytic code will be made available upon reasonable request to the corresponding author. The data are not publicly available due to privacy and/or ethical restrictions.

### Author contributions

*Concept and design*: HKN, KAK and IC; *Acquisition of data and/or lab analysis*: HKN, KAK, RRL and IC; *Statistical analysis*: SA, FK and IC; *Interpretation of data*: All authors; *Drafting the manuscript*: HKN and IC; *Critical revision of manuscript and important intellectual contribution*: All authors; *Obtaining funding*: IC and KAK; *Administrative, technical or material support*: HKN, KAK and IC; *Supervision*: IC; *Primary responsibility for the final content*: IC.

## Abbreviations

BMR_EST_: estimated basal metabolic rate
C-DHQI: Canadian Diet History Questionnaire I
DLW: doubly labeled water
EI: energy intake
NPV: negative predictive values
PAL: physical activity level
PPV: positive predictive value
rEI: self-reported energy intake
STAR-Q: Sedentary Time and Activity Reporting Questionnaire
TEE: total energy expenditure
TBW: total body water

